# The metabolic fingerprint of COVID-19 severity

**DOI:** 10.1101/2020.11.09.20228221

**Authors:** Tim Dierckx, Jan van Elslande, Heli Salmela, Bram Decru, Els Wauters, Jan Gunst, Yannick Van Herck, the CONTAGIOUS-consortium, Joost Wauters, Björn Stessel, Pieter Vermeersch

## Abstract

Corona virus disease 2019 (COVID-19) has been associated with a wide range of divergent pathologies, and risk of severe disease is reported to be increased by a similarly broad range of co-morbidities. The present study investigated blood metabolites in order to elucidate how infection with severe acute respiratory syndrome coronavirus 2 can lead to such a variety of pathologies and what common ground they share. COVID-19 patient blood samples were taken at hospital admission in two Belgian patient cohorts, and a third cohort that included longitudinal samples was used for additional validation (total n=581). A total of 251 blood metabolite measures and ratios were assessed using nuclear magnetic resonance spectroscopy and tested for association to disease severity. In line with the varied effects of severe COVID-19, the range of severity-associated biomarkers was equally broad and included increased inflammatory markers (glycoprotein acetylation), amino acid concentrations (increased leucine and phenylalanine), increased lipoprotein particle concentrations (except those of very low density lipoprotein, VLDL), decreased cholesterol levels (except in large HDL and VLDL), increased triglyceride levels (only in IDL and LDL), fatty acid levels (decreased poly-unsaturated fatty acid, increased mono-unsaturated fatty acid) and decreased choline concentration, with association sizes comparable to those of routine clinical chemistry metrics of acute inflammation. Our results point to systemic metabolic biomarkers for COVID-19 severity that make strong targets for further fundamental research into its pathology (e.g. phenylalanine and omega-6 fatty acids).

## Introduction

Severe corona virus disease 2019 (COVID-19), the disease caused by severe acute respiratory syndrome coronavirus 2 (SARS-CoV-2) infection, is associated with a wide range of pathologies including ARDS^1,2^, lymphocytopenia^3^, acute kidney failure^4^, liver injury^5^ and septic shock^3,4,6^. A similarly wide range of co-morbidities predisposes patients to more severe COVID-19, and includes obesity, diabetics, hypertension and cardiovascular disease (CVD)^7–9^. Studies examining the effects of SARS-CoV-2 infection have implicated a wide variety of immunological processes (reviewed in^10^). SARS-CoV-2 uses the angiotensin-converting enzyme 2 receptor (ACE2) as a cell entry point, which could at least partially explain the diverse effects of SARS-CoV-2 infection, as ACE2 is present on many different cell types. ACE2 has been linked to CVD, inflammation and the metabolism through the renin-angiotensin system (RAS)^11^, and it has been shown to regulate amino acid homeostasis^12^. The populations with increased risk of developing severe COVID-19 share low-grade inflammation as a common feature^13^ and the major risk factor for severe COVID-19 can be interpreted as metabolic dysregulation^14^. A clear need exists for a systemic investigation into the metabolic processes underlying the diverse immunological pathways affected by SARS-CoV-2 to better understand 1) the factors associated with risk of severe infection, 2) the systemic effects of COVID-19, and 3) the cardiometabolic complications following disease resolution.

Limited reports investigating specific metabolites have already been published. These include the association to disease severity of lower levels of low density lipoprotein (LDL)^15^, and cholesterol levels in high density lipoprotein (HDL) and LDL^16^. However, traditional HDL, LDL and cholesterol quantification methods used in clinical settings lack the resolution to analytically capture the lipoprotein profile^17^. Nuclear magnetic resonance (NMR) spectroscopy can be used to address this limitation^18^, and enables a more systemic investigation by simultaneously quantifying a large number of metrics relevant to the COVID-19 setting including energy-related metabolites, amino acid and fatty acid concentrations and glycoprotein acetylation (GlycA)^19,20^.

GlycA is a relatively novel serum biomarker for systemic inflammation, known to be increased in various conditions associated with a predisposition to severe COVID-19, including CVD^21–26^, type-2 diabetes^27^ and elevated BMI^28,29^. The GlycA NMR signal quantifies a non-specific, composite signal that arises from mobile N-acetyl methyl groups of circulating glycoproteins (predominantly alpha-1-acid glycoprotein, alpha-1-antitrypsin, haptoglobin and transferrin)^30^. These glycoproteins are acute-phase reactants, meaning their glycan structures and their concentrations change during the acute-phase response of inflammation^30^. However, GlycA has also shown distinct advantages over traditional inflammatory biomarkers such as C-reactive protein (CRP) in its association to inflammatory conditions^18,31,32^. Due to its more persistent and composite nature, being a read-out for many different inflammatory processes, GlycA can be interpreted as a summary measure of a subject’s ‘baseline’ chronic inflammatory burden^33^. In light of the common theme of low-grade inflammation in populations with higher risk for severe COVID-19^13^ and reports of increased CVD risk during and following COVID-19^34^, GlycA is of particular relevance in the COVID-19 setting.

To elucidate how SARS-CoV-2 infection can lead to such a variety of pathologies and what common ground these pathologies share, we used NMR experiments to quantify chronic inflammatory burden, lipoprotein particle concentrations (as well as their cholesterol and triglyceride content), amino acid and fatty acid concentrations, and energy related metabolites in three sample sets from confirmed SARS-CoV-2 infected patients in two Belgian hospitals, collected at time of hospital admission for COVID-19. We identified biomarkers associated with COVID-19 severity (mild/moderate/severe/critical) and test which of these biomarkers could be used to identify patients who will develop critical disease during their hospital stay. For a limited set of patients, we explore longitudinal data from samples taken at day 7 post admission, at hospital discharge and 30 days post hospital discharge.

## Methods

### Ethics

This study was approved by the Medical Ethical Committee of the University Hospitals of Leuven (MEC, UZ Leuven, s64161 and s63881) and the ethical approval committee of the Jessa hospital (20.75-20.05). Samples from UZL and Jessa cohorts were collected retrospectively, while the CONTAGIOUS cohort study is a prospective observational trial for which informed consent was obtained for all participants (clinicaltrials.gov identifier NCT04327570).

### Study populations

This study included 581 samples from 480 patients across three different cohorts. A graphical overview of the sample sets is available in **Supplementary Figure S1**. Two of the cohorts (UZL, n=219 and JESSA, n=164) consisted of convenience samples of left-over diagnostic samples of unselected adult patients. All COVID-19 patients from the first wave in Belgium (March-June 2020) for whom a left-over sample from routine biochemistry at the time of hospital presentation was available were included. All included patients tested positive for SARS-CoV-2 using real-time polymerase chain reaction, performed in accordance with WHO guidelines^35^. Positive PCR results were obtained at time of admission sampling (median 0 days, range -6 to 2 days for the UZL cohort, range -8 to 0 days for the JESSA cohort, respectively). The UZL cohort consisted of plasma samples, while the samples assayed in the JESSA cohort were serum samples. The third sample set is a subset of plasma samples (n=198, from 97 patients) taken for the CONTAGIOUS observational clinical trial. All consecutive patients (>18 years old) admitted to the University Hospitals of Leuven between March 2020 and September 2020 with PCR-confirmed and/or CT-confirmed SARS-CoV-2 disease (confirmation a median of 2 days prior to admission, range 12 days to 1 day prior to admission) were eligible for inclusion. Samples were taken at the time of admission (within maximum 48h), at day 7, at the time of hospital discharge and 30 days after hospital discharge (if available). Patients for whom admission samples were available within the prospective CONTAGIOUS cohort were excluded from the retrospective UZL cohort.

### Clinical measurements

Severity of infection was retrospectively classified on a scale of 1-4 by a single clinician reviewing individual patient records using the following criteria^36,37^: 1) Mild cases: mild clinical symptoms without manifestation of pneumonia on imaging throughout the duration of disease, 2) Moderate cases: fever, respiratory symptoms, and with radiological findings of pneumonia at any point during the disease, 3) Severe cases: meeting any one of the following criteria during the course of their disease: respiratory distress, hypoxia (SpO_2_ ≤ 93%), or abnormal blood gas analysis (PaO_2_ <60 mmHg, PaCO_2_ >50 mmHg) or 4) Critical cases: meeting any one of the following criteria during the course of their disease: respiratory failure requiring mechanical ventilation, shock, organ failure that requires intensive unit care or death of the patient due to COVID-19.

In the clinical laboratory of the University Hospital Leuven (UZL patient cohort), routine laboratory parameters were measured using Roche Cobas 8000 (Roche, Basel, Switzerland) for clinical chemistry parameters and immunoassays, Sysmex XE 5000 (Sysmex, Kobe, Japan) for hematology parameters and ACL-TOP 700 (Werfen, Milan, Italy) for coagulation tests. In the clinical laboratory of the Jessa hospital, laboratory parameters were quantified using Roche Cobas 8000 (Roche, Basel, Switzerland) for CRP, electrolytes, kidney function, liver function, triglycerides and ferritin, SYSMEX XN-1500 for hematology parameters, and ACL-TOP 550 (Werfen, Milan, Italy) for coagulation tests.

### NMR metabolomics

Nightingale Health Ltd’s (Helsinki, Finland) high-throughput 1H NMR metabolomics platform was used to quantify 251 biomarkers and biomarker ratios. The experimental protocols have been described elsewhere^18,20^. The quantified biomarkers include amino acids, lipoprotein, fatty acids (FA) and triglyceride (TG), in addition to GlycA, creatinine and albumin concentration, as well as ketone bodies and glycolysis related metabolites. The full list of biomarkers quantified by NMR is available in **Supplementary Table S1**. In addition to directly measured metabolic biomarkers, we also included a composite multi-biomarker ‘Infectious Disease Score’ (IDS), calculated as posited by Julkunen et al. (under review^38^). Briefly, this risk score was constructed to associate to the likelihood of severe pneumonia in eight years of follow-up, using NMR measurements taken in biobank samples.

### Statistics

Differences in demographic and clinical measurements between the study populations were assessed using a T-test or Mann-Whitney U test, for normally and non-normally distributed variables respectively, as indicated by a Shapiro-Wilke test for normality. Differences between the datasets of categorical demographic variables were identified using Fisher’s exact test on the contingency tables. Association to severity is reported as odds ratios in univariate cumulative logit ordinal logistic regression on standard deviation (SD) scaled variables. Where the proportional odds assumption was violated, as indicated by the Brant test, the ordinal regression was performed three times, i.e. once for each contrast in the severity scale: contrasting 1) mild versus moderate, severe and critical, 2) mild and moderate versus severe and critical, and finally 3) mild, moderate and severe versus critical. Correction for age and sex had minimal impact on the reported associations (data not shown). Biomarkers with significant association to COVID-19 severity (Benjamini-Hochberg (BH) adjusted p-value<0.05 in both the UZL and Jessa cohorts), were tested for significant (BH adjusted p-value<0.05) differences between severe and critical cases in a third independent cohort (CONTAGIOUS) using a Mann-Whitney U test or Fischer test (for continuous and categorical variables, respectively). All statistical testing was conducted in R v4.0.2^39^ (additional packages: ordinal^40^, brant, ggplot2^41^).

## Results

### Study populations and their differences

We first tested whether there were any significant clinical differences between the two largest patient cohorts (UZL, n=219 and JESSA, n=164, **Table 1**). The demographics of the cohorts were largely comparable, with the exception of slightly higher prevalences of diabetes and CVD in the UZL cohort. Several clinical laboratory measurements taken at admission were also different between the two cohorts: average white blood cell count, neutrophil count and eosinophil count, as well as CRP levels, ferritin levels and activated partial thromboplastin time were significantly higher in the Jessa cohort, whereas slightly higher sodium, potassium and calcium concentrations are noted in the UZL cohort. Some of these differences could be explained by differences in laboratory settings (e.g. UZL quantified total calcium, while Jessa quantified ionized calcium) or patient follow-up (UZL did not consistently quantify ferritin in every sample). We then aimed to identify which biomarkers consistently associated to COVID-19 severity, despite these differences between the study cohorts.

**Table 1:** Demographic and clinical comparison of study populations. Variables with significant differences between the study populations are indicated in bold. CVD: cardiovascular disease, WBC: white blood cell count, aPTT: activated partial thromboplastin time, CRP: c-reactive protein, eGFR: estimated glomerular filtration rate, HBA1c: glycated hemoglobin, GammaGT: gamma-glutamyltransferase, AST: aspartate transaminase, ALT: alanine transaminase.

|  | UZL |  |  |  | Jessa |  |  |  |  |  |
| --- | --- | --- | --- | --- | --- | --- | --- | --- | --- | --- |
| Severity:total samples | Mild: 59 / Moderate: 74<br>Severe: 58 / Critical: 28 |  |  |  | Mild: 37 / Moderate: 48<br>Severe: 51 / Critical: 28 |  |  |  |  |  |
|  | n | Median | IQR |  | n | Median | IQR |  | test | p val |
| Age (years) | 219 | 67 | 56 | 80 | 164 | 72 | 57.75 | 81.25 | Mann-Whitney U | 1.28E-01 |
| BMI (kg/m²) | 154 | 26.5 | 23.3 | 30.0 | 113 | 27.0 | 24.7 | 30.0 | Mann-Whitney U | 2.31E-01 |
| Sex (% female) | 219 | 48% |  |  | 164 | 49% |  |  | Fisher | 8.36E-01 |
| Diabetes (%) | 219 | 27% |  |  | 164 | 15% |  |  | Fisher | 6.05E-03 |
| Hypertension (%) | 219 | 35% |  |  | 164 | 37% |  |  | Fisher | 6.66E-01 |
| CVD (%) | 219 | 35% |  |  | 164 | 19% |  |  | Fisher | 1.17E-03 |
| Chronic Pulmonary Disease (%) | 219 | 16% |  |  | 164 | 18% |  |  | Fisher | 6.80E-01 |
| Hemoglobin (g/dL) | 217 | 13.3 | 12.1 | 14.6 | 163 | 13.3 | 12.05 | 14.3 | Mann-Whitney U | 9.45E-01 |
| WBC (10<sup>9</sup>/L) | 215 | 5.91 | 4.405 | 8.07 | 163 | 6.56 | 4.925 | 8.905 | Mann-Whitney U | 3.98E-02 |
| Neutrophil (10<sup>9</sup>/L) | 207 | 4.1 | 2.85 | 6.3 | 161 | 4.73 | 3.56 | 7.16 | Mann-Whitney U | 1.45E-02 |
| Eosinophil (10<sup>9</sup>/L) | 207 | 0 | 0 | 0 | 160 | 0.005 | 0 | 0.04 | Mann-Whitney U | 4.13E-08 |
| Lymphocyte (10<sup>9</sup>/L) | 208 | 1 | 0.7 | 1.4 | 161 | 0.88 | 0.54 | 1.43 | Mann-Whitney U | 9.69E-02 |
| Platelets (10<sup>9</sup>/L) | 216 | 195 | 154 | 266.3 | 163 | 205 | 163 | 269.5 | Mann-Whitney U | 3.73E-01 |
| APTT (s) | 145 | 27.8 | 26 | 31.3 | 150 | 33.7 | 30.2 | 39 | Mann-Whitney U | 1.85E-16 |
| Fibrinogen (g/L) | 38 | 4.02 | 3.235 | 5.225 | 5 | 6.06 | 4.2 | 7.39 | Mann-Whitney U | 9.18E-02 |
| D-dimer (µg/L) | 70 | 624.5 | 442 | 1197 | 144 | 780.5 | 515.8 | 1388 | Mann-Whitney U | 1.04E-01 |
| CRP (mg/L) | 213 | 40.8 | 11.1 | 84.4 | 162.0 | 72.0 | 19.3 | 120.0 | Mann-Whitney U | 2.89E-03 |
| Sodium (mmol/L) | 213 | 138.5 | 135.4 | 140.3 | 162 | 137 | 135 | 139 | Mann-Whitney U | 1.39E-02 |
| Potassium (mmol/L) | 214 | 4.04 | 3.705 | 4.28 | 162 | 3.89 | 3.633 | 4.26 | Mann-Whitney U | 1.52E-02 |
| Creatinine (mg/dL) | 215 | 0.95 | 0.73 | 1.22 | 163 | 0.95 | 0.78 | 1.25 | Mann-Whitney U | 2.34E-01 |
| eGFR (mL/min/1.73 m²) | 213 | 74 | 57 | 94 | 163 | 72 | 50 | 86 | Mann-Whitney U | 1.09E-01 |
| Calcium (mmol/L) | 186 | 2.29 | 2.213 | 2.34 | 76 | 1.14 | 1.11 | 1.18 | Mann-Whitney U | 5.80E-37 |
| Glucose (mg/dL) | 193 | 115 | 101 | 138 | 139 | 119 | 106 | 140.5 | Mann-Whitney U | 2.45E-01 |
| HbA1c (% ratio) | 116 | 6.05 | 5.8 | 6.525 | 6 | 6.1 | 5.925 | 7.1 | Mann-Whitney U | 7.09E-01 |
| Total Bilirubin (mg/dL) | 198 | 0.465 | 0.34 | 0.618 | 158 | 0.51 | 0.38 | 0.7 | Mann-Whitney U | 8.94E-02 |
| Alkaline Phosphatase (U/L) | 210 | 70 | 54 | 92 | 154 | 67.5 | 55 | 90.5 | Mann-Whitney U | 6.34E-01 |
| Gamma GT (U/L) | 211 | 40 | 24 | 71.5 | 160 | 42 | 24 | 76 | Mann-Whitney U | 6.29E-01 |
| AST (U/L) | 213 | 32 | 24 | 52 | 161 | 34 | 24 | 47 | Mann-Whitney U | 9.79E-01 |
| ALT (U/L) | 214 | 24 | 16 | 37 | 161 | 25 | 17 | 38 | Mann-Whitney U | 2.75E-01 |
| Creatine Kinase (U/L) | 133 | 86 | 59 | 192 | 3 | 178 | 118.5 | 178 | Mann-Whitney U | 6.73E-01 |
| Troponin (µg/L) | 141 | 0.016 | 0.008 | 0.033 | 147 | 0.0158 | 0.008 | 0.029 | Mann-Whitney U | 7.17E-01 |
| Ferritin (µg/L) | 66 | 327.5 | 121.1 | 1026 | 144 | 710 | 337.5 | 1400 | Mann-Whitney U | 1.44E-04 |

### Identifying metabolic biomarkers for COVID-19 severity

Using ordinal regression on sd-scaled clinical and NMR measurements, 130 biomarkers were found to be significantly associated to disease severity in both the UZL and Jessa patient cohorts (full results in **Supplementary Table S2**). At admission, only eight of the measured demographic and clinical biomarkers were found to be significantly associated to COVID-19 severity in both patient cohorts (age, CVD status, O_2_ saturation, creatinine and eGFR, CRP, and lymphocyte and eosinophil count) (**Figure 1**). Some demographic parameters expected to associate to severity based on previous reports (e.g. sex, BMI and diabetic status), trended towards association but fell short of statistical significance in both cohorts simultaneously.

**Figure 1:**
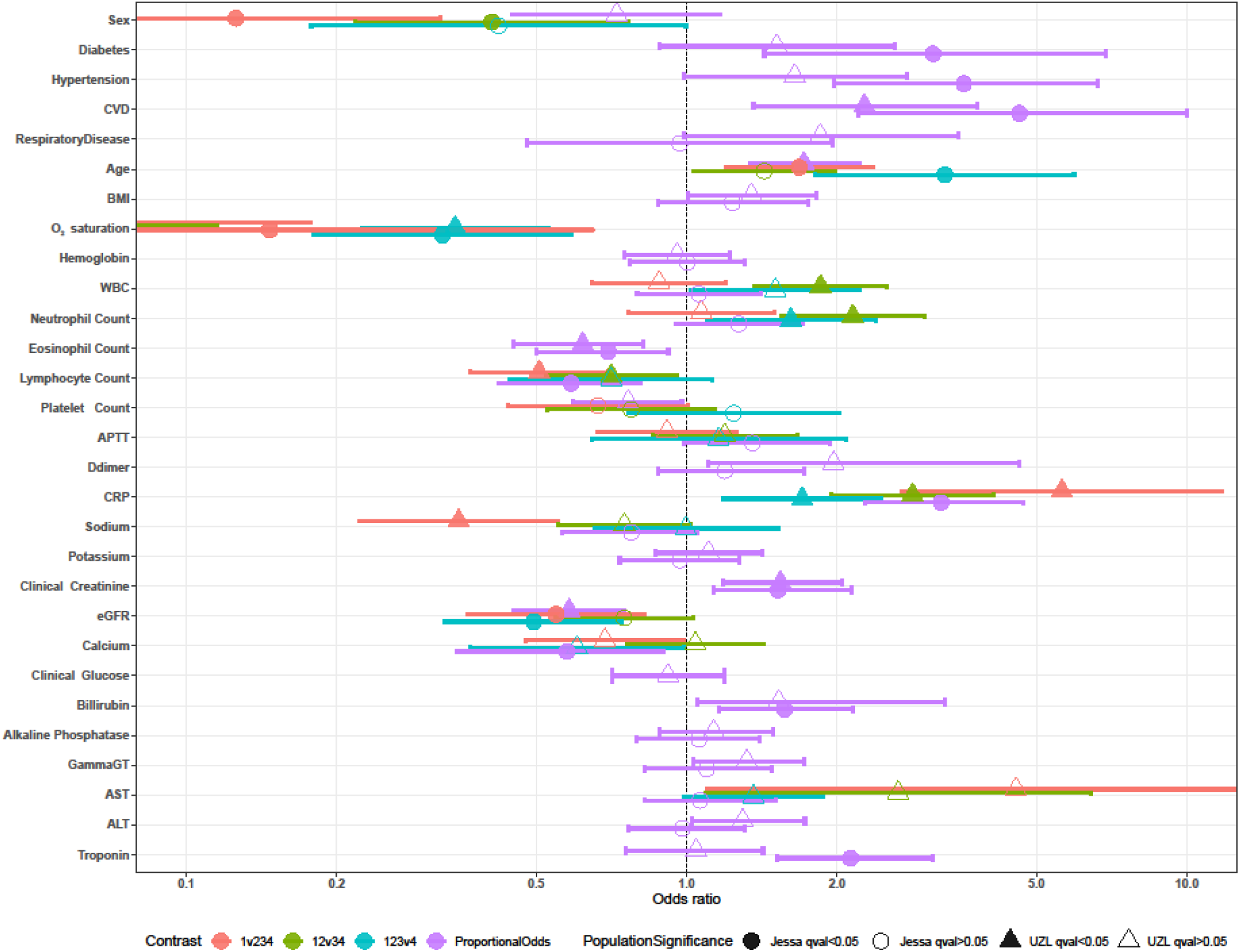
Odds Ratios of clinical and demographic biomarkers associations with COVID-19 severity. Association of demographic and clinical variables with disease severity in the Jessa (n=164, circle) and UZL cohort (n=219, triangle) tested using ordinal regression on sd-scaled variables. Odds ratios of proportional odds in each cohort are presented in purple. If odds ratios differ significantly between each severity class contrast, as judged by a Brant test, the OR is visualized separately for each contrast: red: Mild (severity 1) versus Non-mild (severity 2, 3 and 4), green: Mild and Moderate (severity 1 and 2) versus Severe and Critical (severity 3 and 4), and finally, blue: non-critical (severity 1, 2 and 3) versus critical (severity 4). Multiple-comparison corrected significance of each association is indicated by the opacity of each point (opaque: q-value < 0.05, clear: q-value > 0.05). ALT: alanine aminotransferase, APTT: activated partial thromboplastin time, AST: aspartate transaminase, BMI: body mass index, CRP: c-reactive protein, CVD: cardiovascular disease, eGFR: estimated glomerular filtration rate, WBC: white blood cell count.

Of the 251 metabolites and metabolite ratios quantified by NMR, 122 associated with COVID-19 severity in both cohorts. In line with the clinical associations, NMR evidence of inflammation (GlycA, albumin, and the GlycA/albumin ratio) and kidney function (creatinine) showed strong associations with COVID-19 severity (**Figure 2**). These metrics are important components of Julkunen et al’s ‘Infectious Disease Score’ (IDS), which can partly explain the IDS’s strong association to COVID-19 severity (odds ratios of 4 and 4.5 for the UZL and Jessa cohort, respectively). Of the 9 quantified amino acids, phenylalanine (Phe) and leucine (Leu) show consistent association to severity. Valine and total branched-chain amino acid concentrations were associated to severity in the UZL cohort, but not the Jessa cohort. Other notable associations only observed in a single cohort include glutamine (Gln), tyrosine (Tyr) and Histidine (His) (**Figure 3**).

**Figure 2:**
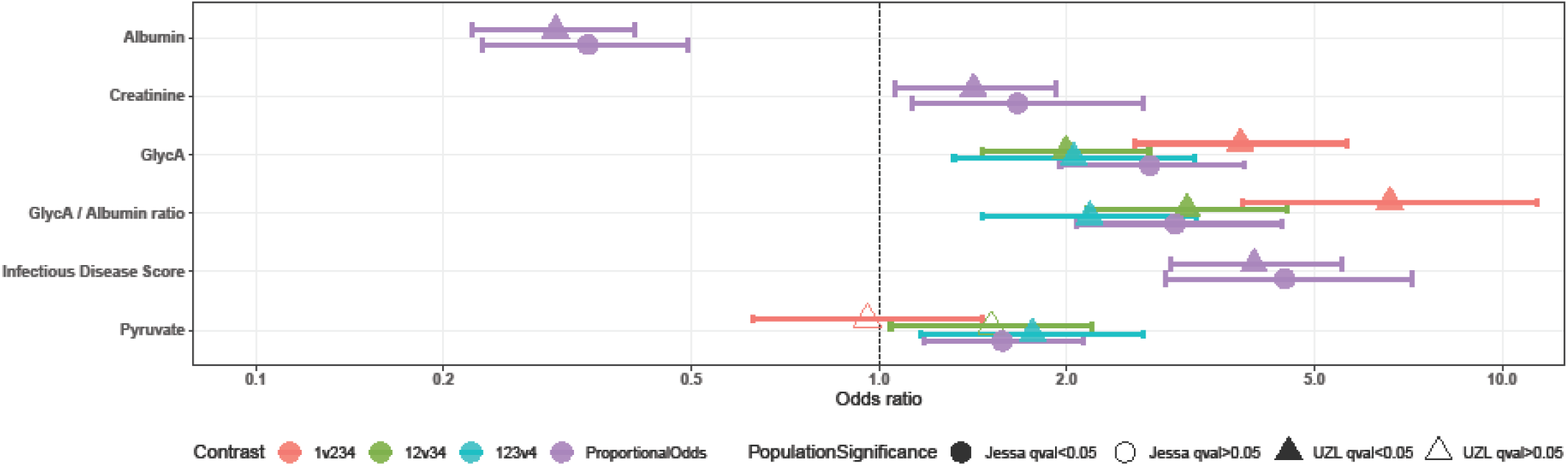
Odds ratios of Inflammation-related NMR biomarker associations with COVID-19 severity. In the Jessa (n=164, circle) and UZL cohort (n=219, triangle), associations of NMR biomarkers were tested using ordinal regression on sd-scaled variables. Odds ratios of proportional odds in each cohort are presented in purple. If odds ratios differ significantly between each severity class contrast, as judged by a Brant test, the OR is visualized separately for each contrast: red: Mild (severity 1) versus Non-mild (severity 2, 3 and 4), green: Mild and Moderate (severity 1 and 2) versus Severe and Critical (severity 3 and 4), and finally, blue: non-critical (severity 1, 2 and 3) versus critical (severity 4). Multiple-comparison corrected significance of each association is indicated by the opacity of each point (opaque: q-value < 0.05, clear: q-value > 0.05). GlycA: Glycoprotein Acetylation.

**Figure 3:**
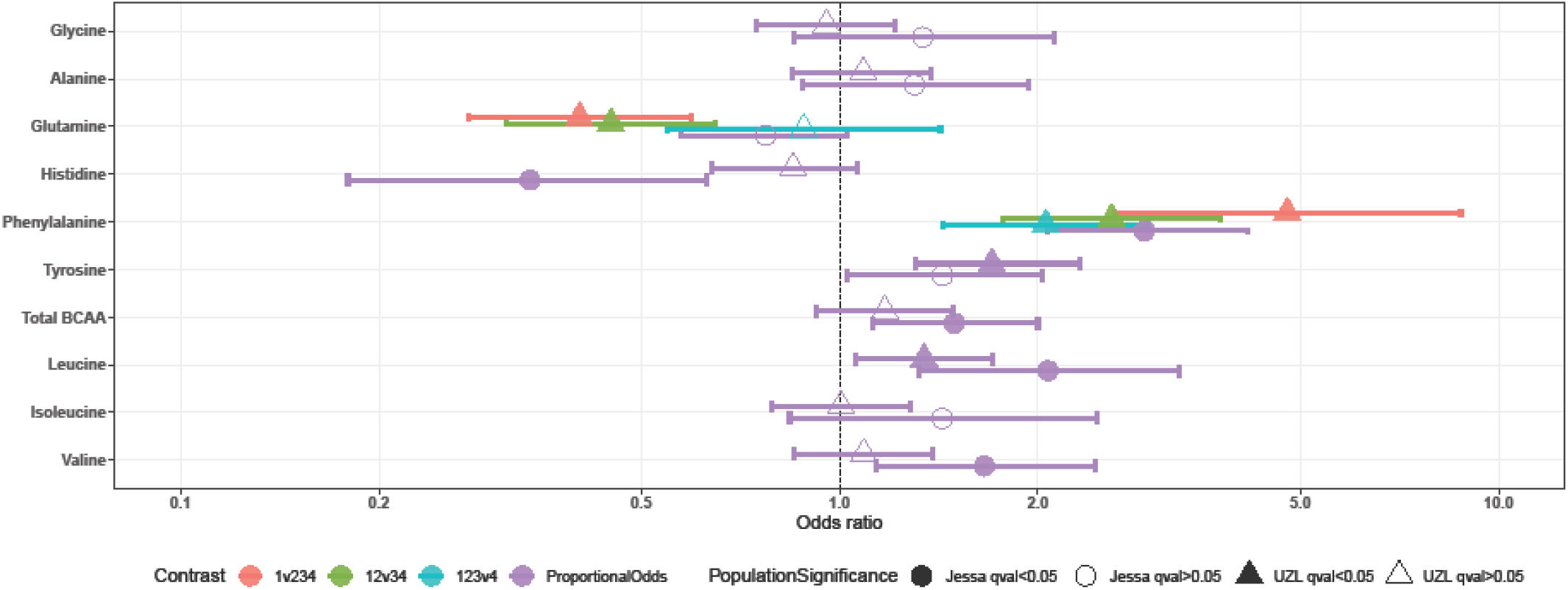
Odds ratios of amino-acid concentration associations with COVID-19 severity. In the Jessa (n=164, circle) and UZL cohort (n=219, triangle), associations of NMR biomarkers were tested using ordinal regression on sd-scaled variables. Odds ratios of proportional odds in each cohort are presented in purple. If odds ratios differ significantly between each severity class contrast, as judged by a Brant test, the OR is visualized separately for each contrast: red: Mild (severity 1) versus Non-mild (severity 2, 3 and 4), green: Mild and Moderate (severity 1 and 2) versus Severe and Critical (severity 3 and 4), and finally, blue: non-critical (severity 1, 2 and 3) versus critical (severity 4). Multiple-comparison corrected significance of each association is indicated by the opacity of each point (opaque: q-value < 0.05, clear: q-value > 0.05). BCAA: branched-chain amino acids.

Concentrations and content of analytical lipoprotein subclasses associated well with COVID-19 severity. Overall, higher lipoprotein concentrations and higher cholesterol content was associated with less severe disease, with the notable exception of VLDL particles and of the larger HDL particles. Higher triglyceride content of lipoprotein particles was associated with disease severity, but only for IDL and LDL (**Figure 4**). While increased absolute phospholipid content in HDL, IDL and LDL is associated with less severe disease, this contrasted to the relative phospholipid to total lipid ratios in HDL, IDL and LDL, which were associated with higher disease severity. Relative lipoprotein content ratios (including relative cholesterol, cholesteryl ester, free cholesterol, and triglyceride to total lipid ratios) otherwise showed similar association to severity as their absolute concentrations (**Supplementary Table S2**).

**Figure 4:**
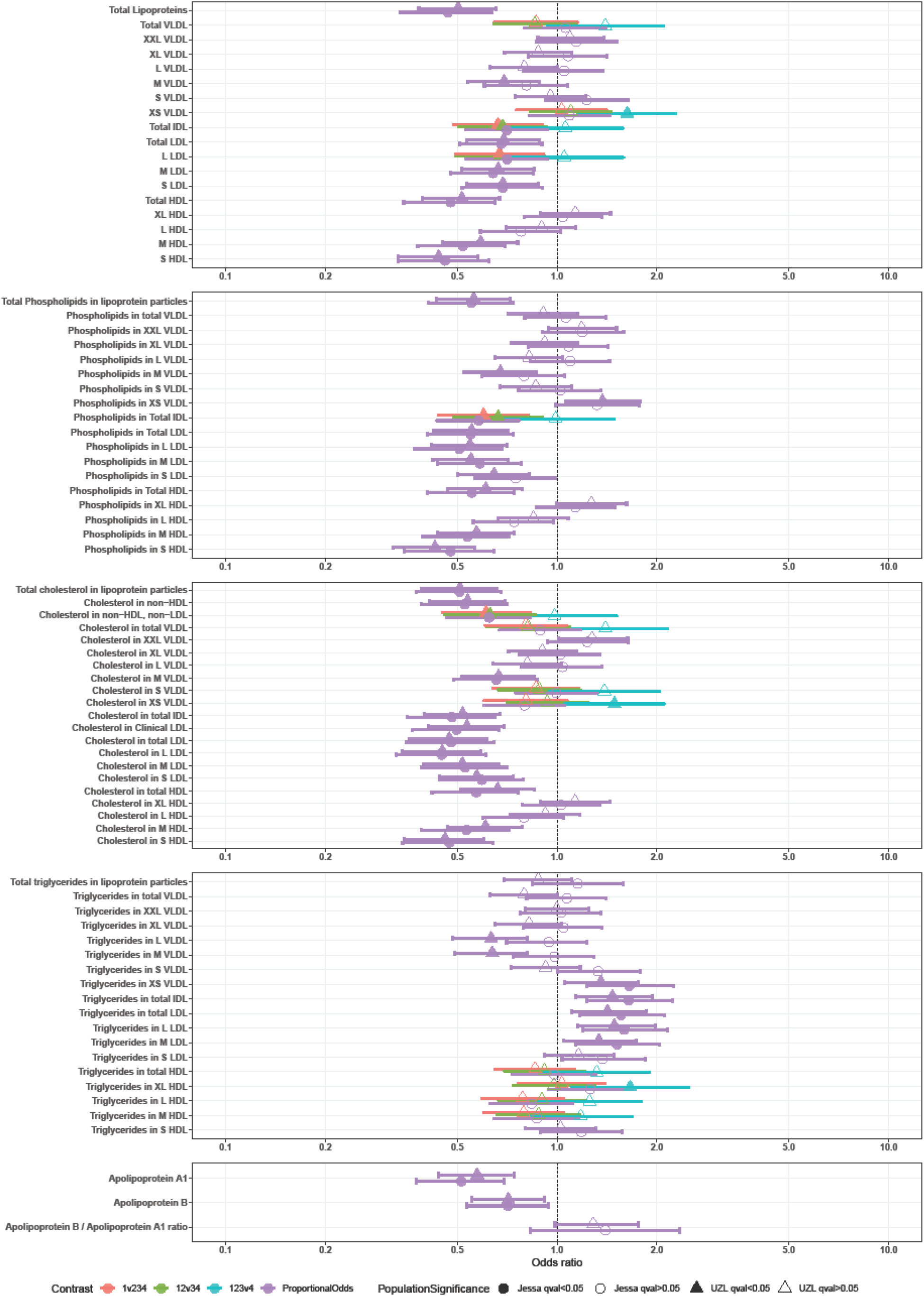
Odds ratios of lipoprotein particle and lipoprotein particle content associations with COVID-19 severity. In the Jessa (n=164, circle) and UZL cohort (n=219, triangle), associations of NMR biomarkers were tested using ordinal regression on sd-scaled variables. Odds ratios of proportional odds in each cohort are presented in purple. If odds ratios differ significantly between each severity class contrast, as judged by a Brant test, the OR is visualized separately for each contrast: red: Mild (severity 1) versus Non-mild (severity 2, 3 and 4), green: Mild and Moderate (severity 1 and 2) versus Severe and Critical (severity 3 and 4), and finally, blue: non-critical (severity 1, 2 and 3) versus critical (severity 4). Multiple-comparison corrected significance of each association is indicated by the opacity of each point (opaque: q-value < 0.05, clear: q-value > 0.05). VLDL: very-low-density lipoprotein, IDL: intermediate-density lipoprotein, LDL: low-density lipoprotein, HDL: high-density lipoprotein. Lipoprotein subclass sizes are listed in **Supplementary Table S1**.

Quantification of FA showed increased poly-unsaturated FA (PUFA) content was associated with less severe disease, while increased mono-unsaturated FA (MUFA) content was associated to more severe disease (**Figure 5**). Linoleic acid (LA) and total Omega-6 FA showed stronger and more consistent associations with COVID-19 severity than Omega-3 FA. Both total Omega-3 FA and docosahexaenoic acid (DHA) showed opposite associations in the two examined cohorts (OR 0.72 and 0.80 in UZL, OR 1.2 and 1.4 in Jessa, for Omega-3 FA and DHA ratios to total FA, respectively).

**Figure 5:**
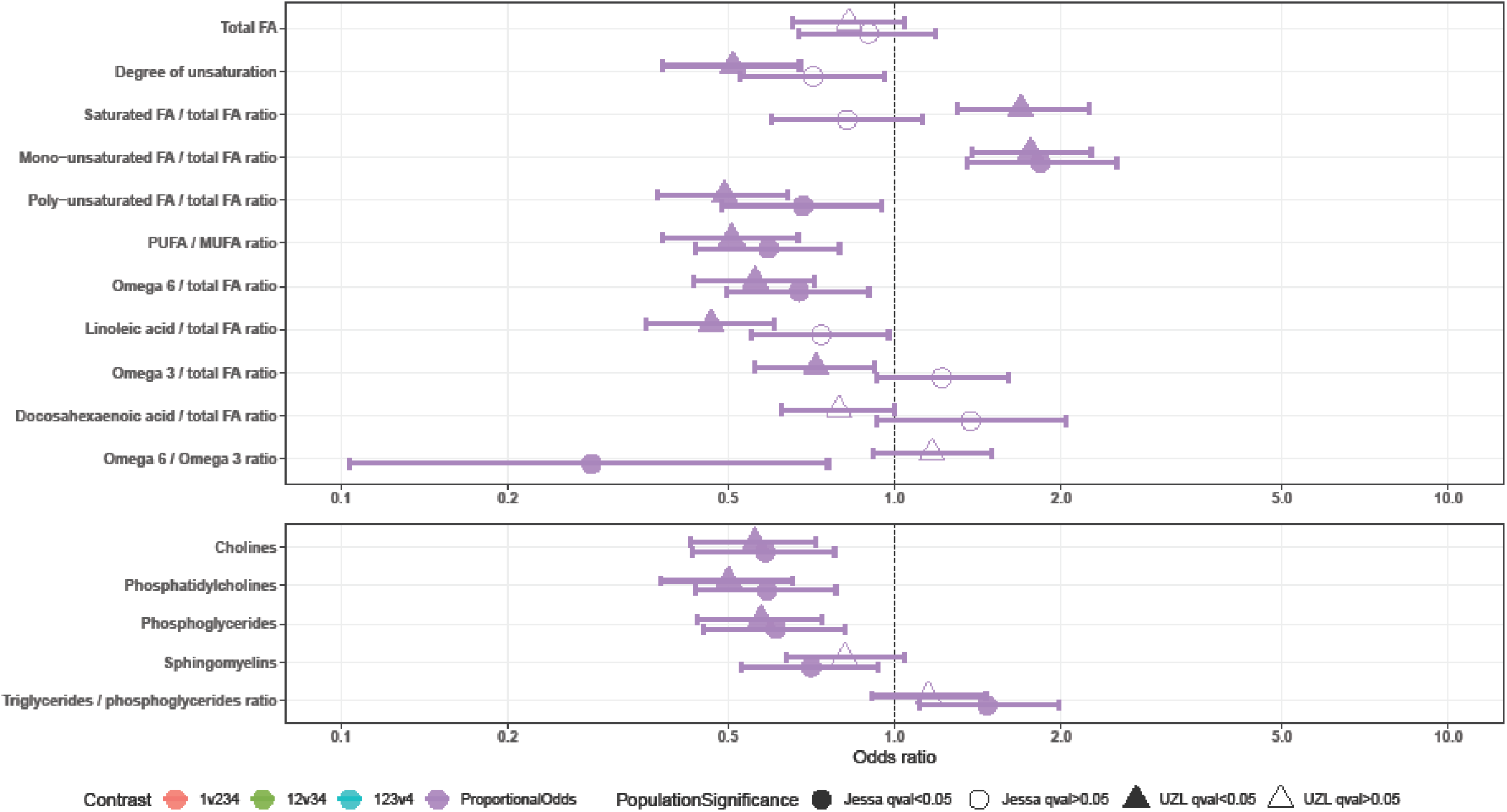
Odds ratios of COVID-19 severity associations of fatty acids and lipid-associated metabolites. In the Jessa (n=164, circle) and UZL cohort (n=219, triangle), associations of NMR biomarkers were tested using ordinal regression on sd-scaled variables. Odds ratios of proportional odds in each cohort are presented in purple. If odds ratios differ significantly between each severity class contrast, as judged by a Brant test, the OR is visualized separately for each contrast: red: Mild (severity 1) versus Non-mild (severity 2, 3 and 4), green: Mild and Moderate (severity 1 and 2) versus Severe and Critical (severity 3 and 4), and finally, blue: non-critical (severity 1, 2 and 3) versus critical (severity 4). Multiple-comparison corrected significance of each association is indicated by the opacity of each point (opaque: q-value < 0.05, clear: q-value > 0.05). FA: Fatty Acid. MUFA: mono-unsaturated FA, PUFA: poly-unsaturated FA.

### Validation in CONTAGIOUS cohort

To further validate our results, we tested which of the 130 biomarkers that were consistently associated with severity showed significant differences between severe and critical disease severity classes in admission samples (n=72) from a third, independent, patient cohort (CONTAGIOUS, clinical and demographic parameters at admission, comparisons to UZL and Jessa cohorts, **Supplementary Table S3**). In total, 72 severity-associated biomarkers (5 clinical, 68 NMR) showed statistically significant differences at admission between patients with severe and critical disease (n=48 and n=24, respectively), with strong correlations between some of the quantified moieties (**Figure 6**). Selected biomarkers are presented in **Figure 7:** albumin, GlycA/albumin ratio and IDS retained their association to disease in the CONTAGIOUS setting that does not include patients with mild or moderate disease. Phenylalanine was the only amino acid that retained its significance in this context. Omega-6 FA, PUFA and PUFA/MUFA ratio findings were consistent with our earlier observations. Again, HDL cholesterol content was significantly lower at admission in the more severe COVID-19 cases, and this difference was only evident in the small HDL sizes.

**Figure 6:**
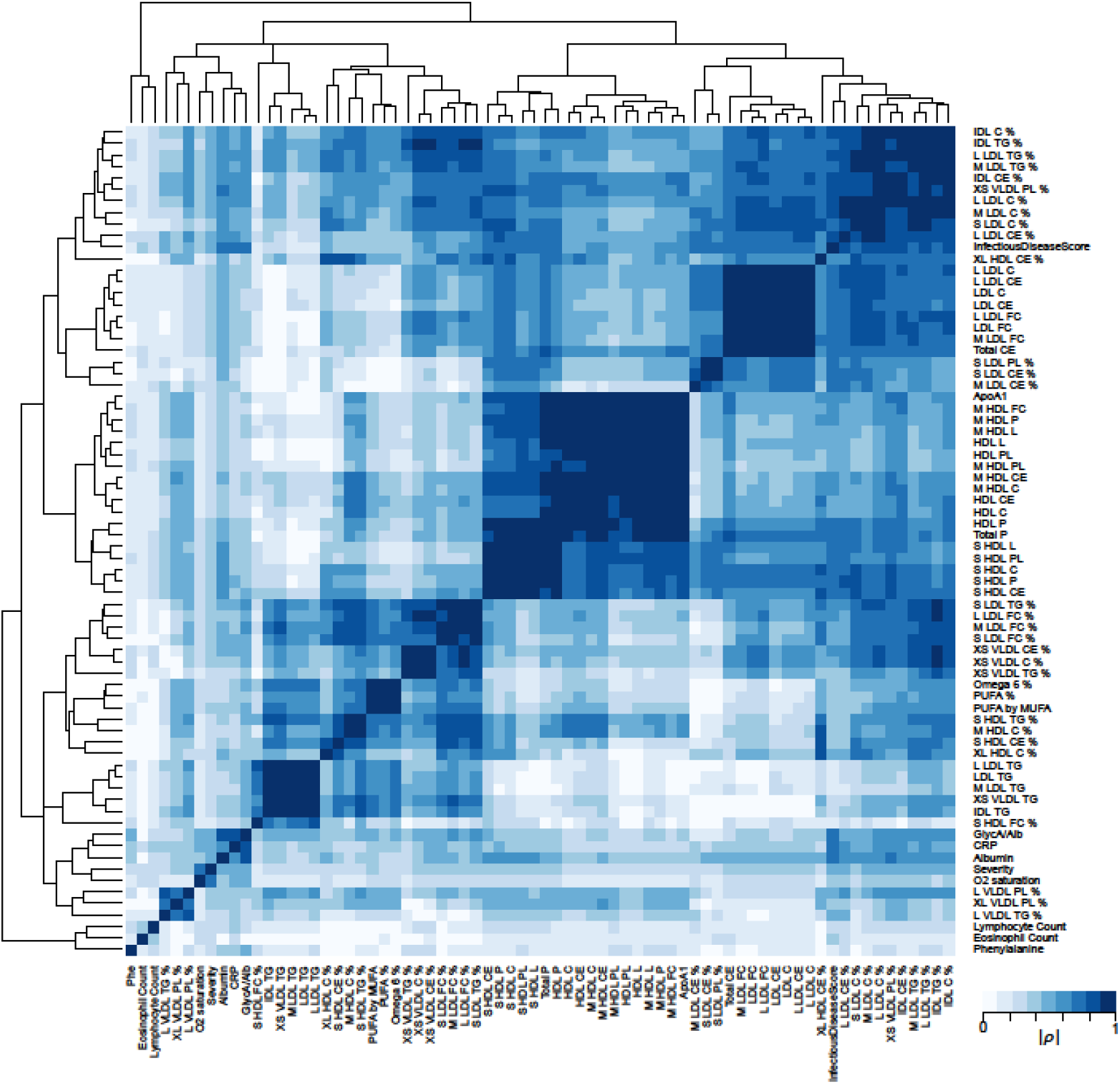
Hierarchical clustering of Spearman correlations of COVID-19 severity associated biomarkers. All depicted biomarkers are significantly associated to severity in both the UZL and Jessa cohorts and show significant differences between severe and critical cases of the CONTAGIOUS patient cohort. Hierarchical clustering was achieved by average linkage of the absolute value of Spearman’s rho. C: cholesterol, CE cholesteryl ester, FC, free cholesterol, TG: triglyceride, FA: fatty acid, L: total lipid concentration, P: lipoprotein particle concentration, PL: phospholipid concentration, PUFA poly-unsaturated FA, MUFA: mono-unsaturated FA, VLDL: very-low-density lipoprotein, IDL: intermediate-density lipoprotein, LDL: low-density lipoprotein, HDL: high-density lipoprotein. Lipoprotein subclass sizes are listed in **Supplementary Table S1**.

**Figure 7:**
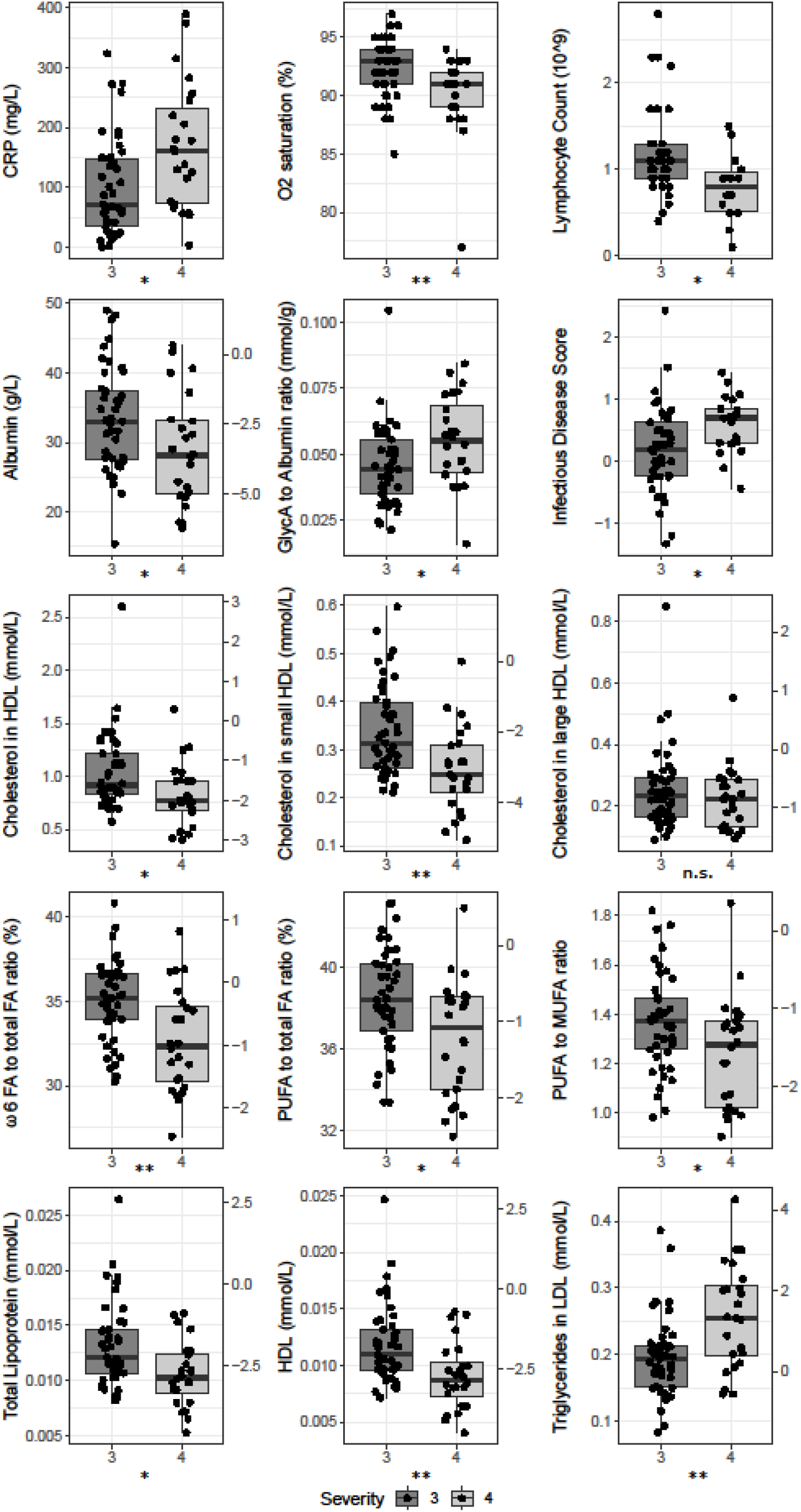
Selected biomarkers in severe and critical cases of the CONTAGIOUS cohort. Severity 3 and 4, severe and critical, respectively. Where depicted, the y-axis on the right side of the graph normalizes the data using the mean and standard deviations observed in Finnish demographic cohorts^42^. n.s. not significant, * p<0.05, ** p<0.01. CRP: c-reactive protein, FA: fatty acid, HDL: high-density lipoprotein, LDL, low-density lipoprotein.

### Longitudinal associations

For the CONTAGIOUS cohort of severe and critically afflicted patients (n=97), longitudinal samples were available taken at day 7 post admission (D7, n=38), at hospital discharge (Dis, n=41), and 30 days post discharge (Dis+30, n=47). Compared to samples taken at admission, most severity-associated biomarkers worsened at D7, and return to admission levels at time of hospital discharge (full results in **Supplementary Table S4**). At 30 days post discharge, clinical biomarkers for severity (CRP, Ferritin) returned to healthy levels (**Figure 8**). Likewise, GlycA as well as the GlycA/albumin ratio dropped below admission levels by day 30 post hospital discharge and returned to concentrations observed in healthy controls reported in demographic studies^38,42^. Choline, phosphatidylcholine and phosphoglyceride concentrations, which were significantly associated to severity (**Figure 5**), showed no change during the first seven days of hospitalization and up until discharge, but were found to be significantly increased 30 days post discharge.

**Figure 8:**
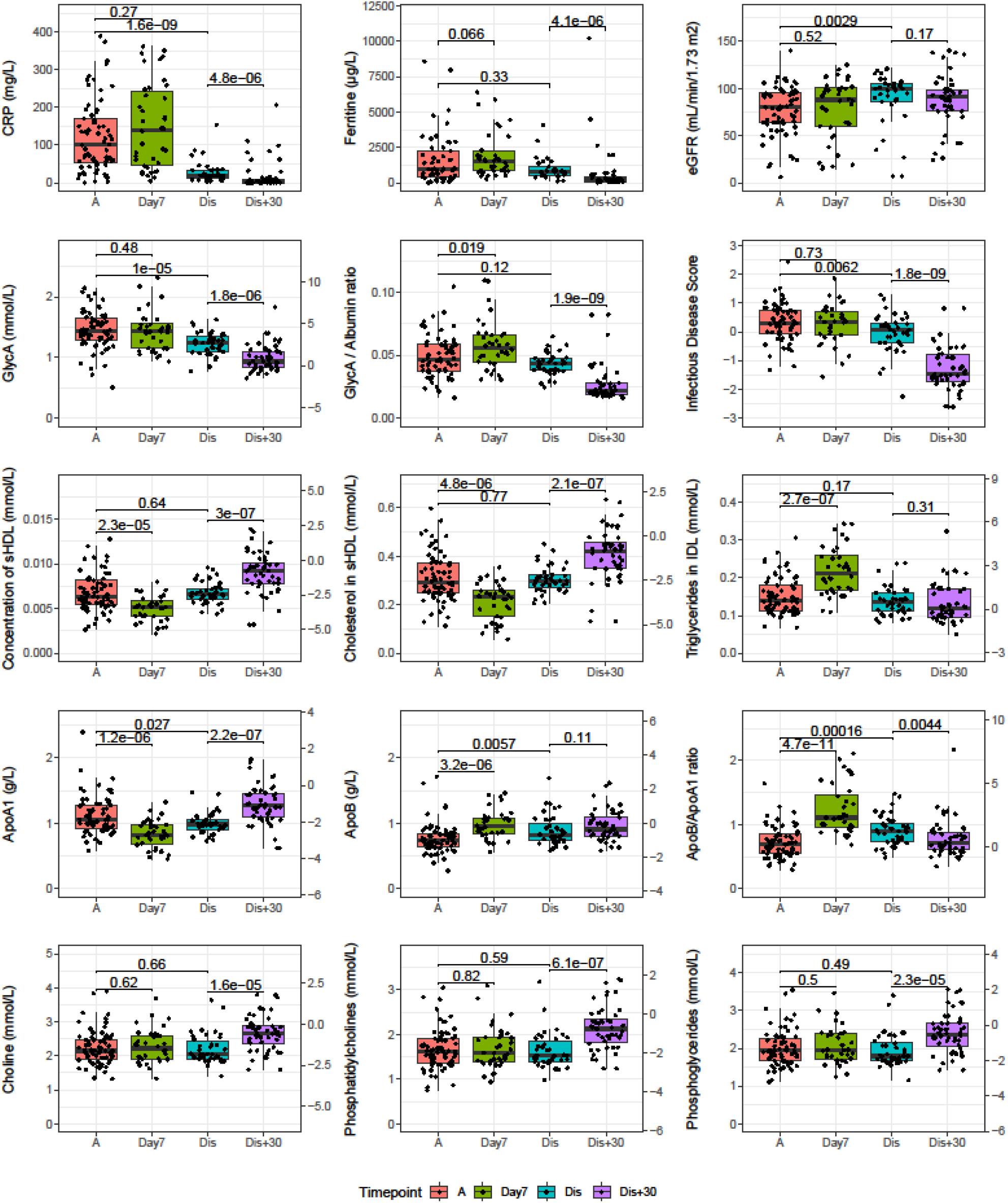
Longitudinal analysis of severe and critical COVID patients of the CONTAGIOUS cohort. Patients of the CONTAGIOUS cohort were sampled at COVID admission (A, red), 7 days post admission (Day7, green), time of hospital discharge (Dis, blue) and 30 days post hospital discharge (Dis+30, purple). Indicated are p-values of Mann-Whitney U tests comparing the indicated timepoints. Where depicted, the y-axis on the right side of the graph normalizes the data using the mean and standard deviations observed in Finnish demographic cohorts^45^. Full comparison results for all variables associated to severity are available in **supplementary table S4**. ApoA1: Apolipoprotein A1, ApoB: Apolipoprotein B, CRP: c-reactive protein, GlycA: glycoprotein acetylation.

## Discussion

We performed the first in-depth characterization of circulating metabolites in SARS-CoV-2 infected patients across the spectrum of disease severity. In left-over diagnostic samples taken at time of hospital admission of two independent cohorts (n=219 and n=164), we identified a metabolic profile associated to COVID-19 severity. This metabolic fingerprint of COVID-19 is relevant to four aspects of COVID-19 related research: first, we provide further metabolic evidence of the association between COVID-19 severity and inflammation. Second, our amino acid profiling confirms observations of altered energy requirements of hyperinflammatory immune cells of COVID-19 patients^43,44^ and suggests an alternative explanation for the low occurrence of severe COVID-19 cases in phenylketonuria patients^45^. Third, we refine existing reports on the association between lipoprotein particles, their cholesterol content, and COVID-19 severity^15,16^ by investigating size-specific lipoprotein subclasses and their content. Finally, we add to the literature relating COVID-19 severity to fatty acid concentrations in the blood, and observe an association with linoleic acid that is in-line with mechanistic evidence^46^.

Metabolic evidence of inflammation, i.e. increased GlycA levels and reduced albumin levels, was associated with increased severity (**Figure 2**). Many populations reported as high-risk for developing severe COVID-19^7^ share low-grade chronic inflammation as a common feature^13^. Increased GlycA concentrations have been reported in populations with co-morbidities related to increased risk of severe COVID-19^7,9^ such as advanced age^24^, diabetes^47^, obesity^48^, and CVD^21–23,33^. GlycA association to severity is further strengthened by calculating the GlycA to Albumin ratio (**Figure 2**). While routine clinical chemistry metrics of acute inflammation such as CRP and ferritin showed stronger associations to severity than GlycA (**Figure 1**), GlycA has higher basal healthy control levels^38,42^, higher longitudinal stability compared to CRP^49^ and is associated with inflammatory disease severity even in CRP negative patients^32^, which could be of when investigating the longitudinal effects of COVID-19 (**Figure 8**).

In line with reports on reduced mitochondrial oxidative phosphorylation in hyperinflammatory immune cells^43^ of COVID-19 patients^44^, our results confirmed higher serum pyruvate concentration to be associated with increased COVID-19 severity (**Figure 2**). The observed associations of amino-acid concentrations with severity could be interpreted in a similar light (**Figure 3**): the lower serum glutamine (GLN) concentration in severe COVID-19 could represent increased GLN catabolism in visceral tissues to accommodate the energy requirements of hyperinflammatory cells^50^ through anaplerosis. Our findings suggest that in mild cases, ongoing branched chain amino acid (BCAA) oxidation contributes to sufficient availability of GLN, while in severe cases BCAAs accumulate in serum as their oxidation is inhibited in favor of cytosolic aerobic glycolysis. Other than leucine, phenylalanine also showed a consistently significant association with COVID-19 severity (with borderline significance observed for tyrosine, **Figure 3**). Phenylalanine’s association to severity is in line with reports of low occurrence of severe COVID-19 outcomes among phenylketonuria patients, which was previously ascribed to this population’s lower rates of vitamin D deficiency as a result of the protein substitutes in their diet^45^.

Our results confirm reports of low lipoprotein particle concentrations of LDL^15^, HDL, and their cholesterol content^16^ in severe COVID-19. However, these early reports were limited both in the scope of the considered lipoprotein particles (only examining HDL and LDL) and in the degree to which these particles are characterized (only quantifying their cholesterol content). The 1H NMR panel assayed in the present study comprehensively characterizes the range of lipoprotein particle densities and sizes, and provides an in-depth profile of each of the particle subclasses. We expand on existing observations to show that not all HDL sizes associate to severity: very large and large HDL particles, and their lipid and cholesterol content, showed no significant association to severity (**Figure 4**), while small HDL was among the strongest and most consistent associations to COVID-19 severity. Decreased small HDL has been reported to be associated with higher all-cause mortality risk^51^, though these findings contrast to associations noted in CVD studies. In the CVD context, large HDL is inversely associated to CVD risk, while small HDL typically shows no such association^52^. Similarly opposite to what is reported in the CVD context, our findings show increased LDL concentration, as well as its phospholipid and cholesterol content, is associated with decreased severity. Triglyceride associations to COVID-19 severity are similar to what’s observed in CVD and diabetes^53^: high TG levels in IDL, LDL and VLDL and low TG levels in HDL, though this latter association does not reach significance in our cohorts (**Figure 4**).

While the altered cholesterol profile of severe COVID-19 can be interpreted as metabolic evidence of inflammation, it can also be construed as evidence of a genetic predisposition to severe infection, similar to what’s been shown for HDL cholestrol^54^. Relevant to the hyperinflammatory state^13^ frequently identified as a “cytokine storm” in COVID-19^55^, a meta-analysis of genome-wide association studies has shown a genetic link between cytokine activity and the serum concentration of apolipoprotein B, serum cholesterol and the cholesterol and phospholipid content of IDL, LDL and VLDL^56^, all of which are shown to be lower in severe COVID-19 cases in our results. This interpretation in terms of a predisposition to severe infection finds additional support in the observed association at admission between severity and the blood biomarker score for infectious disease risk, which was constructed by Julkunen et al. from biobank samples to associate to the likelihood of severe pneumonia and severe COVID-19 during eight years of follow-up^38^.

Higher relative PUFA content and PUFA to MUFA ratio were consistently associated with lower severity, in contrast to increased MUFA levels (**Figure 5**). Linoleic Acid concentration and total omega-6 fatty acid content (absolute concentration and relative to total FA content) were lower in severe COVID-19 cases in our cohorts. Conflicting reports exist on PUFA association with inflammation, and omega-6 FA have been linked to increased inflammation in the airways^57^. Considering a recent report showed that linoleic acid (a PU omega-6 FA) tightly binds to the spike glycoprotein which is used by SARS-CoV-2 to bind the ACE2 receptor to gain entry into host cells^46^, the association between less severe COVID-19 and higher omega-6 PUFA levels could therefore be a mechanistic effect, rather than an effect mediated through inflammation.

Interestingly, one recent report showed that the precursor for phosphatidylcholine, phosphocholine, was upregulated in severe COVID-19^58^, while our results show increased phosphatidylcholine is associated with less severe COVID-19, as are serum phosphoglycerides and choline levels, in agreement with other findings^58^ (**Figure 5**). Furthermore, analysis of longitudinal follow-up samples of severe and critical patients showed no changes in these concentrations during hospitalization (**Figure 8**), but a marked increase of these concentrations was observed in samples taken 30 days after hospital discharge. This, combined with the convenience with which the 1H NMR spectrum can be measured and how many of the captured biomarkers are relevant to COVID-19 severity, leads us to suggest that the NMR analysis technique is well-suited to prospective longitudinal studies aiming to elucidate the long-term effects of COVID-19.

We point out several strengths and limitations of our study. The studied populations had significant differences in demographics and clinical characteristics, and differences in sample type (serum vs plasma) between our two largest cohorts discourage the merging of their experimental results, even after accounting for the batch effects introduced by differences in sample handling between the different cohorts. However, this does lend a high degree of confidence to the results that were consistently observed in all three cohorts. Availability of longitudinal follow-up samples was limited to the CONTAGIOUS cohort of patients with severe infection. It is worth noting that, to our knowledge, this is the first study using 1H NMR metabolomics in severe viral infection. As such, without comparable results from studies on other infections, we cannot attribute the results reported in this study exclusively to SARS-CoV-2 infection. We suspect that e.g. a study into the metabolic associations of severe pneumonia could yield similar results, in light of the strength of the association of severity with the infectious disease multi-biomarker score from Julkunen et al^38^. Many of the associations to severity noted here could be interpreted as general signs of inflammation and robust correlation network analysis on paired NMR metabolomics, cytokine quantification and transcriptomics, while contrasting severe COVID-19 to severe (viral) pneumonia without SARS-CoV-2 infection, would allow for a more fine-grained interpretation of the results.

COVID-19 is reported to have many diverse effects and complications, many of which can be related to low-grade inflammation. Our study of systemic metabolic biomarkers reveals that the metabolic fingerprint of COVID-19 severity consists of a pervasive inflammatory signature, not unlike reported associations with CVD risk or long-term risk of pneumonia. The broad range of biomarkers reflects the wide range of COVID-19 symptoms and co-morbidities, and these biomarkers should be further investigated for their translational potential, and their precise role in the COVID-19 pathology.

## Data Availability

For purposes of reproducing the results and additional research, data can be made available from the authors upon request.

## Acknowledgements

The authors thank Kimberly Vanhees, Cato Jacobs, Evy Smeyers and Laurien Geebelen and for their efforts in sample selection and preparation, and the procurement of clinical data.

## References

1. Gattinoni, L., Chiumello, D. & Rossi, S. COVID-19 pneumonia: ARDS or not? Crit. Care 24, 154 (2020).

2. Li, X. & Ma, X. Acute respiratory failure in COVID-19: is it “typical” ARDS? Crit. Care 24, 198 (2020).

3. Bhatraju, P. K. et al. Covid-19 in Critically Ill Patients in the Seattle Region — Case Series. N. Engl. J. Med. NEJMoa2004500 (2020). doi:10.1056/NEJMoa2004500

4. Arentz, M. et al. Characteristics and Outcomes of 21 Critically Ill Patients With COVID-19 in Washington State. JAMA 4720, 2019–2021 (2020).

5. Cai, Q. et al. COVID-19: Abnormal liver function tests. J. Hepatol. 73, 566–574 (2020).

6. Chen, T. et al. Clinical characteristics of 113 deceased patients with coronavirus disease 2019: retrospective study. BMJ 2, m1091 (2020).

7. Yang, J. et al. Prevalence of comorbidities and its effects in patients infected with SARS-CoV-2: a systematic review and meta-analysis. Int. J. Infect. Dis. 94, 91–95 (2020).

8. Richardson, S. et al. Presenting Characteristics, Comorbidities, and Outcomes Among 5700 Patients Hospitalized With COVID-19 in the New York City Area. JAMA 323, 2052 (2020).

9. Hamer, M., Gale, C. R., Kivimäki, M. & Batty, G. D. Overweight, obesity, and risk of hospitalization for COVID-19: A community-based cohort study of adults in the United Kingdom. Proc. Natl. Acad. Sci. 117, 21011–21013 (2020).

10. Sokolowska, M. et al. Immunology of COVID-19: mechanisms, clinical outcome, diagnostics and perspectives – a report of the European Academy of Allergy and Clinical Immunology (EAACI). Allergy all.14462 (2020). doi:10.1111/all.14462

11. Gheblawi, M. et al. Angiotensin-Converting Enzyme 2: SARS-CoV-2 Receptor and Regulator of the Renin-Angiotensin System. Circ. Res. 126, 1456–1474 (2020).

12. Hashimoto, T. et al. ACE2 links amino acid malnutrition to microbial ecology and intestinal inflammation. Nature 487, 477–481 (2012).

13. Ciornei, R. T. Prevention of Severe Coronavirus Disease 2019 Outcomes by Reducing Low-Grade Inflammation in High-Risk Categories. Front. Immunol. 11, 1–5 (2020).

14. Ayres, J. S. A metabolic handbook for the COVID-19 pandemic. Nat. Metab. 2, 572– 585 (2020).

15. Fan, J. et al. Letter to the Editor: Low-density lipoprotein is a potential predictor of poor prognosis in patients with coronavirus disease 2019. Metabolism 107, 154243 (2020).

16. Wei, X. et al. Hypolipidemia is associated with the severity of COVID-19. J. Clin. Lipidol. 14, 297–304 (2020).

17. Holmes, M. V. & Ala-Korpela, M. What is ‘LDL cholesterol’? Nat. Rev. Cardiol. 16, 197– 198 (2019).

18. Soininen, P., Kangas, A. J., Würtz, P., Suna, T. & Ala-Korpela, M. Quantitative Serum Nuclear Magnetic Resonance Metabolomics in Cardiovascular Epidemiology and Genetics. Circ. Cardiovasc. Genet. 8, 192–206 (2015).

19. Wishart, D. S. NMR metabolomics: A look ahead. J. Magn. Reson. 306, 155–161 (2019).

20. Soininen, P. et al. High-throughput serum NMR metabonomics for cost-effective holistic studies on systemic metabolism. Analyst 134, 1781 (2009).

21. Gruppen, E. G. et al. GlycA, a Pro-Inflammatory Glycoprotein Biomarker, and Incident Cardiovascular Disease: Relationship with C-Reactive Protein and Renal Function. PLoS One 10, e0139057 (2015).

22. Connelly, M. A., Otvos, J. D., Shalaurova, I., Playford, M. P. & Mehta, N. N. GlycA, a novel biomarker of systemic inflammation and cardiovascular disease risk. J. Transl. Med. 15, 219 (2017).

23. Joshi, A. A. et al. GlycA Is a Novel Biomarker of Inflammation and Subclinical Cardiovascular Disease in Psoriasis. Circ. Res. 119, 1242–1253 (2016).

24. Akinkuolie, A. O., Buring, J. E., Ridker, P. M. & Mora, S. A Novel Protein Glycan Biomarker and Future Cardiovascular Disease Events. J. Am. Heart Assoc. 3, e001221– e001221 (2014).

25. Wurtz, P. et al. Metabolite Profiling and Cardiovascular Event Risk: A Prospective Study of 3 Population-Based Cohorts. Circulation 131, 774–785 (2015).

26. Ballout, R. A. & Remaley, A. T. GlycA: a new biomarker for systemic inflammation and cardiovascular disease (CVD) risk assessment. J. Lab. Precis. Med. 5, 17–17 (2020).

27. Dullaart, R. P. F., Gruppen, E. G., Connelly, M. A. & Lefrandt, J. D. A pro-inflammatory glycoprotein biomarker is associated with lower bilirubin in metabolic syndrome. Clin. Biochem. 48, 1045–1047 (2015).

28. Würtz, P. et al. Metabolic Signatures of Adiposity in Young Adults: Mendelian Randomization Analysis and Effects of Weight Change. PLoS Med. 11, e1001765 (2014).

29. Dullaart, R. P. F., Gruppen, E. G., Connelly, M. A., Otvos, J. D. & Lefrandt, J. D. GlycA, a biomarker of inflammatory glycoproteins, is more closely related to the leptin/adiponectin ratio than to glucose tolerance status. Clin. Biochem. 48, 811–814 (2015).

30. Bell, J. D., Brown, J. C. C., Nicholson, J. K. & Sadler, P. J. Assignment of resonances for ‘acute-phase’ glycoproteins in high resolution proton NMR spectra of human blood plasma. FEBS Lett. 215, 311–315 (1987).

31. Otvos, J. D. et al. GlycA: A Composite Nuclear Magnetic Resonance Biomarker of Systemic Inflammation. Clin. Chem. 61, 714–723 (2015).

32. Dierckx, T., Verstockt, B., Vermeire, S. & van Weyenbergh, J. GlycA, a nuclear magnetic resonance spectroscopy measure for protein glycosylation, is a viable biomarker for disease activity in IBD. J. Crohns. Colitis 1–6 (2018). doi:10.1093/ecco-jcc/jjy162

33. Lawler, P. R. et al. Circulating N-Linked Glycoprotein Acetyls and Longitudinal Mortality RiskNovelty and Significance. Circ. Res. 118, 1106–1115 (2016).

34. Long, B., Brady, W. J., Koyfman, A. & Gottlieb, M. Cardiovascular complications in COVID-19. Am. J. Emerg. Med. 38, 1504–1507 (2020).

35. Corman, V. M. et al. Detection of 2019 novel coronavirus (2019-nCoV) by real-time RT-PCR. Eurosurveillance 25, 1–8 (2020).

36. Qu, J. et al. Profile of Immunoglobulin G and IgM Antibodies Against Severe Acute Respiratory Syndrome Coronavirus 2 (SARS-CoV-2). Clin. Infect. Dis. 10–13 (2020). doi:10.1093/cid/ciaa489

37. Van Elslande, J. et al. Antibody response against SARS-CoV-2 spike protein and nucleoprotein evaluated by four automated immunoassays and three ELISAs. Clin. Microbiol. Infect. (2020). doi:10.1016/j.cmi.2020.07.038

38. Julkunen, H. et al. Blood biomarker score identifies individuals at high risk for severe COVID-19 a decade prior to diagnosis metabolic profiling of 105000 adults in the UK Biobank. Medrxiv 1–15 (2020). doi:10.1101/2020.07.02.20143685

39. R Core Team. R: A Language and Environment for Statistical Computing. (2017).

40. Christensen, R. H. B. ordinal—Regression Models for Ordinal Data. (2019).

41. Wickam, H. ggplot2: Elegant Graphics for Data Analysis. (Springer-Verlag New York, 2009).

42. Tikkanen, E. et al. Metabolic Biomarkers for Peripheral Artery Disease Compared with Coronary Artery Disease. medRxiv 1–16 (2020). doi:10.1101/2020.07.24.20158675

43. Bar-Or, D. et al. Overcoming the Warburg Effect: Is it the key to survival in sepsis? J. Crit. Care 43, 197–201 (2018).

44. Reiter, R. J. et al. Melatonin Inhibits COVID-19-induced Cytokine Storm by Reversing Aerobic Glycolysis in Immune Cells: A Mechanistic Analysis. Med. Drug Discov. 6, 100044 (2020).

45. Rocha, J. C., Calhau, C. & MacDonald, A. Reply to Jakovac; Severity of COVID-19 infection in patients with phenylketonuria: is vitamin D status protective? Am. J. Physiol. Metab. 318, E890–E891 (2020).

46. Toelzer, C. et al. Free fatty acid binding pocket in the locked structure of SARS-CoV-2 spike protein. Science (80-.). 3255, eabd3255 (2020).

47. Connelly, M. A. et al. GlycA, a marker of acute phase glycoproteins, and the risk of incident type 2 diabetes mellitus: PREVEND study. Clin. Chim. Acta 452, 10–17 (2016).

48. Manmadhan, A. et al. Elevated GlycA in severe obesity is normalized by bariatric surgery. Diabetes. Obes. Metab. 21, 178–182 (2019).

49. Connelly, M. A. et al. Differences in GlycA and lipoprotein particle parameters may help distinguish acute kawasaki disease from other febrile illnesses in children. BMC Pediatr. 16, 151 (2016).

50. Holeček, M. Branched-chain amino acids in health and disease: metabolism, alterations in blood plasma, and as supplements. Nutr. Metab. (Lond). 15, 33 (2018).

51. Deelen, J. et al. A metabolic profile of all-cause mortality risk identified in an observational study of 44,168 individuals. Nat. Commun. 10, 3346 (2019).

52. Holmes, M. V. et al. Lipids, Lipoproteins, and Metabolites and Risk of Myocardial Infarction and Stroke. J. Am. Coll. Cardiol. 71, 620–632 (2018).

53. Krauss, R. M. Lipids and Lipoproteins in Patients With Type 2 Diabetes. Diabetes Care 27, 1496–1504 (2004).

54. Madsen, C. M., Varbo, A., Tybjærg-Hansen, A., Frikke-Schmidt, R. & Nordestgaard, B. G. U-shaped relationship of HDL and risk of infectious disease: two prospective population-based cohort studies. Eur. Heart J. 39, 1181–1190 (2018).

55. Mehta, P. et al. COVID-19: consider cytokine storm syndromes and immunosuppression. Lancet 395, 1033–1034 (2020).

56. Nath, A. P. et al. Multivariate Genome-wide Association Analysis of a Cytokine Network Reveals Variants with Widespread Immune, Haematological, and Cardiometabolic Pleiotropy. Am. J. Hum. Genet. 105, 1076–1090 (2019).

57. Rutting, S. et al. Dietary omega-6, but not omega-3, polyunsaturated or saturated fatty acids increase inflammation in primary lung mesenchymal cells. Am. J. Physiol. Cell. Mol. Physiol. 314, L922–L935 (2018).

58. Shen, B. et al. Proteomic and Metabolomic Characterization of COVID-19 Patient Sera. Cell 182, 59-72.e15 (2020).

