## Supplementary material for "The metabolic fingerprint of COVID-19 severity": Figure S1

28 clinical & demographic
251 NMR biomarkers

→

8 clinical & demographic
122 NMR biomarkers

→

5 clinical & demographic
67 NMR biomarkers

Test for association to severity in two cohorts

Significant difference between
severe and critical in third cohort?

UZ (219 patients, 219 samples)

At COVID Admission: 59/74/58/28

**Plasma** sample remainders of diagnostic samples

Jessa (164 patients, 164 samples)

At COVID Admission: 37/48/51/28

**Serum** sample remainders of diagnostic samples

CONTAGIOUS (97 patients, 198 samples)

At COVID Admission: 0/0/48/24
At Day 7 follow-up: 0/0/8/30
At Discharge: 0/0/15/26
At 30d follow-up: 0/0/25/22

Plasma samples, structured collection

Severity at COVID admission: Mild/Moderate/Severe/Critical
